# The CT Scan Lung Severity Score and Vaccination Status in COVID-19 patients in India: Perspective of an Independent Radiology Practice

**DOI:** 10.1101/2021.07.15.21260597

**Authors:** Revat T. Lakhia, Jaimin R. Trivedi

**Author notes:** Correspondence Jaimin R. Trivedi, MBBS, MPH, Assistant Professor, Department of Cardiovascular and Thoracic Surgery, University of Louisville School of Medicine, 201 Abraham Flexner Way, Suite 1200, Louisville, KY 40202, (708) 606 – 3115.

## Abstract

**Background:** The COVID-19 patients often undergo a high-resolution CT scan to determine extent of their lung involvement. The aim of this study was to determine lung involvement in confirmed/suspected COVID-19 patients (encountered at an independent radiology practice) and its correlation to vaccination status amidst the second COVID-19 wave in India.

**Methods:** We retrospectively queried our data since April 2021 to identify adult patients (>17 years) who had confirmed (positive RT-PCR or antigen test) or suspected COVID-19 (classic symptoms but negative RT-PCR) and received a high-resolution CT scan to determine the extent of their lung involvement using the CT severity (CT-SS) score. Patients were classified in 3 groups based on their vaccination status to determine its correlation with the CT-SS score: fully vaccinated, partially vaccinated, and unvaccinated. Basic descriptive statistics, univariate tests and multivariate linear regression analysis were used.

**Results:** We identified 229 patients (median age 45 years, 60% male) of which 205 (89%) had confirmed COVID-19 (positive RT-PCR) and 24 had suspected disease (negative RT-PCT but classic symptoms). Of the 229 patients 29 (13%) had complete vaccination, 38 (17%) had partial vaccination and 162 (70%) had no vaccination. The CT score of the completely vaccinated patients was significantly lower compared to partially or unvaccinated patients (median 0 v. 3.5 v. 10, respectively p<.01). A multivariate linear regression model showed that partial or fully vaccinated patients had lower CT severity score compared to unvaccinated patients (adjusted R squared = 0.47).

**Conclusion:** We present here the real-world findings from an independent radiology practice (a unique and common practice model), in India amid the second COVID-19 wave showing significantly lower CT severity score in fully or partially vaccinated patients compared to unvaccinated patients. Complete vaccination in patients could be critical in preventing severe lung disease.

## Introduction

The novel coronavirus (SARS nCOV-2) impacted over 160 million people across the globe causing severe respiratory illness in many patients with mortality of over 2%^1^. Countries like USA, UK and South Africa have seen deadlier second waves due to highly infectious mutated strains; one such (B1617 or delta) is currently rampant in India crippling the already constrained healthcare infrastructure^2^. There are some patients with symptomatic disease but have negative RT-PCR or rapid antigen test. These patients often undergo a high-resolution CT scan to determine extent of their lung involvement and aid in further management^3^.

Unlike the western countries, the radiology practice in India is largely done by independently operated groups (without affiliations to hospitals or care centers).^4^ A common patient is often referred to these independent practices including suspected/confirmed COVID-19 patients to determine the degree of their lung involvement. We have seen several patients in our practice during the second COVID-19 wave who had negative RT-PCR test but presented with classic symptoms such as fever, fatigue, body aches, occasional shortness of breath etc. Our case mix included fully or partially vaccinated or completely unvaccinated patients. At the time of the study there were 2 vaccines approved in India, both of which required 2 doses for complete vaccination^5^. The aim of this study was to determine lung involvement in confirmed/suspected COVID-19 patients and its correlation to vaccination status.

## Methods

We received an approval from an independent ethical committee from our hometown in Ahmedabad, India. The ACEAS independent ethical committee reviewed our protocol an approved our study. A request of waiver of informed consent also approved considering the retrospective nature of the study. We retrospectively queried our data since April 2021 to identify adult patients (>17 years) who had confirmed (positive RT-PCR or antigen test) or suspected COVID-19 (classic symptoms but negative RT-PCR) and received a high-resolution CT scan to determine the extent of their lung involvement using the CT severity score (CT-SS)^6^. Along with the RT-PCR results, we recorded vaccination status at the time of their CT scan. Patients with both doses completed 1 week prior to CT scan were considered fully vaccinated whereas patients receiving CT scan 2 weeks after the 1st dose or within 1 week of 2nd dose were considered partially vaccinated and the rest of the patients (not receiving any vaccine or within 2 weeks of 1st dose) were considered unvaccinated^7^. Based on the vaccination age eligibility criteria we stratified the cohort in 3 groups of 18-44 years, 45-59 years, and 60 years or above.

We used basic descriptive statistics (median (interquartile range) and % (N)), univariate tests (non-parametric (Kruskal-Wallis) test for continuous variables and chi-square test for categorical variables) and multivariate linear regression analysis to determine the association of the CT-SS score to RT-PCR and vaccination status. All the analyses were conducted using SAS 9.4 software (SAS Inc. Cary, NC) at 95% confidence interval level.

There was no public or patient participation in design of this study.

## Results

We identified 229 patients of which 205 (89%) had confirmed COVID-19 (positive RT-PCR) and 24 had suspected disease (negative RT-PCT but classic symptoms). The median age of the cohort was 45 (36-58) years, and 138 (60%) patients were male. The median CT score of the entire cohort was 9 (2-13). There were 49% (n=112), 29% (n=67) and 22% (n=50) patients in age groups 18-44 years, 45-59 years and > 60 years, respectively. Median CT score in patients in age group 18-44 years was 9 (0-12), 45-60 years was 11 (6-15) and >60 years was 8 (1-11).

Of the 229 patients only 29 (13%) had complete vaccination, 38 (17%) had partial vaccination and 162 (70%) had no vaccination. Of the RT-PCR positive patients, 29 (14%) had complete vaccination, 32 (15%) had partial vaccination. The median age in the vaccination groups was comparable (median 42 v. 48 v. 43, p=0.32, Table 1). There were more male patients in the unvaccinated group compared to fully or partially vaccinated groups (67% v. 42% v. 47%, p<.01). The overall CT score in RT-PCR positive cohort was significantly higher compared to the RT-PCR negative (median 10 vs. 0, p<.01) cohort. The CT score of the completely vaccinated patients was significantly lower compared to the partially or unvaccinated patients (median 0 v. 3.5 v. 10, p<.01, Figure 1). Within RT-CPR positive cohort, patients with complete vaccination had significantly lower CT-SS score compared to partially or unvaccinated patients (median 0 vs. 4 vs. 11, p=0.02).

**Table 1.** Baseline characteristics of patients stratified by vaccination status

|  | Fully Vaccinated<br>N=29 | Partially Vaccinated<br>N=38 | Not vaccinated<br>N=162 | p-value |
| --- | --- | --- | --- | --- |
| Age (years) | 42 (32-64) | 48 (40-56) | 43 (36-57) | 0.32 |
| Gender Male | 42% (12) | 47% (18) | 67% (108) | <.01 |
| RT-PCR positive | 100% (29) | 84% (32) | 89% (144) | 0.10 |
| Median CT score | 0 (0-3) | 3.5 (0-9) | 10 (7-14) | <.01 |
| Data presented as median (interquartile range), % (n). |  |  |  |  |

**Figure 1.**
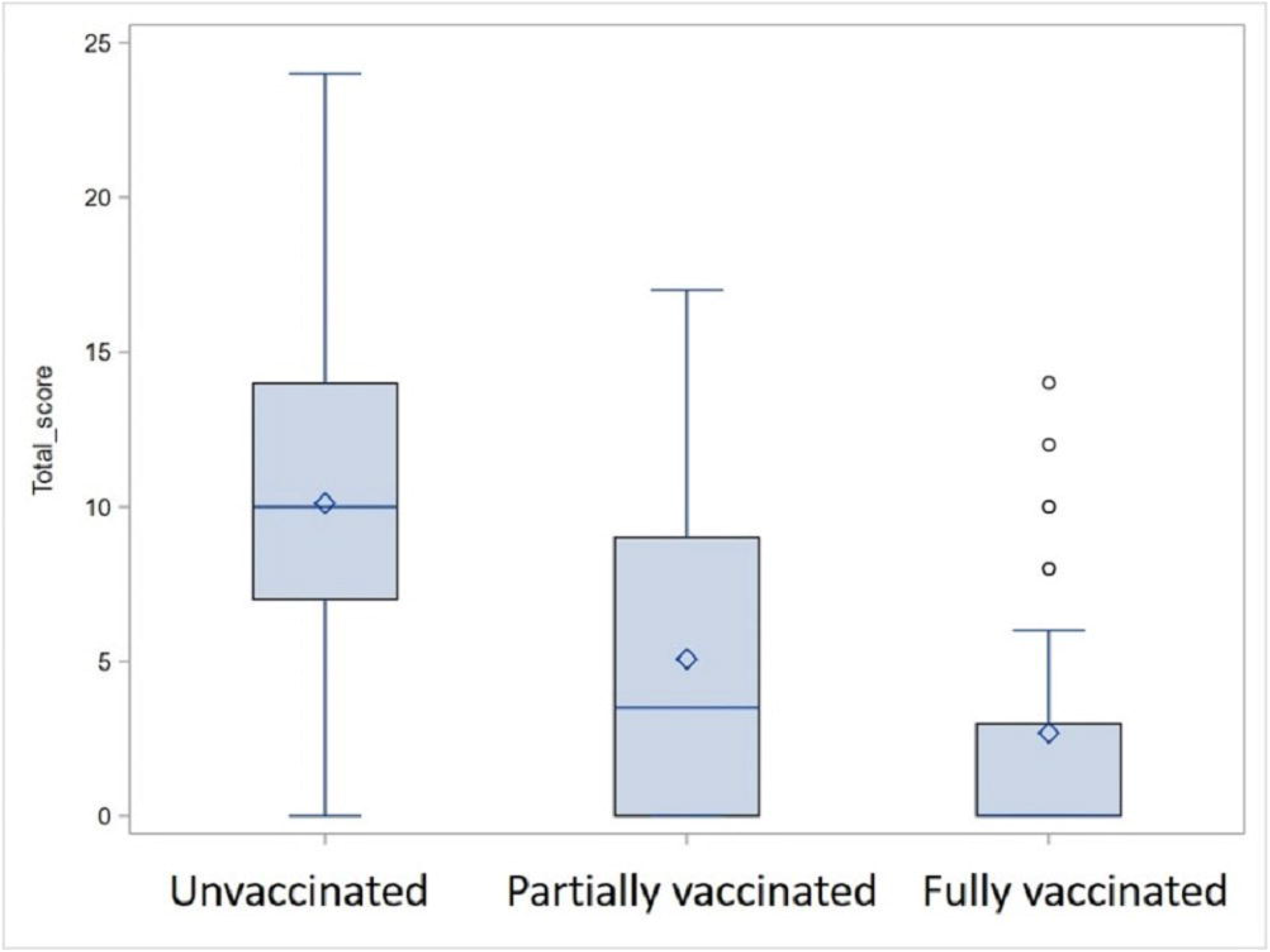
Median CT severity (CT-SS) score stratified by vaccination status

A multivariate linear regression model showed that higher age and presence of positive RT-PCR test were associated with higher CT severity score and partial or fully vaccinated patients had lower CT severity score compared to unvaccinated patients (adjusted R squared = 0.47).

## Discussion

The second COVID-19 wave has put extreme pressure on India’s healthcare system anecdotally affecting younger patients^2^. We observed that almost half of all the patients in our cohort were 45 years or younger which is correlated with then national vaccination guidelines (vaccination for healthcare and frontline workers and aged over 45 years starting). We also observed that patients with complete or even partial vaccination had lower CT severity score which reattributes the effectiveness of the vaccines in preventing the severe disease^7^. In the 11% RT-PCR negative patients, we observed a low CT severity score (median 0). This summary describes COVID-19 patient profiles and its correlation with the CT-SS score from an independent radiology practice perspective in India.

Even though radiology practice in India is predominantly independent, they likely encounter majority of COVID-19 cases needing imaging tests, including severely ill patients who may eventually get admitted to the hospitals for further treatment^4 8^. Our study represents a unique and common practice across India and report the real-world evidence of the CT scan findings and its association with vaccination status and RT-PCR positivity in COVID-19 patients.

Vaccine effectiveness has been scrutinized with emergence of SARS nCOV-2 variants as B1617 (delta) or B117 (alpha) mutants appear to have higher transmissibility and case fatality than the unmutated version of the virus^9 10^. Previous studies have suggested that vaccines are effective against variants in preventing severe disease^11 12^. A recent report from India suggested that fully vaccinated patients may still get infected with the delta (B1617) variant however there was no mortality and high-grade fever was predominant symptom^13^. Similarly, our study showed that 29 fully vaccinated patients had a positive RT-PCR test. Although genomic sequencing data in our cohort was unavailable, based on the previous study, delta variant (B1617) could be predominantly responsible as our patients are from the same timeframe and geography.

Our study shows that compared to unvaccinated patients, completely or partially vaccinated patients had significantly lower CT severity score, irrespective of their RT-PCR status which corroborates the evidence that vaccines are particularly effective tool in preventing a severe COVID-19 even if effectiveness in limiting the spread could be lower in leu of emerging variants^7 12 13^. This finding of reduced lung involvement with full or partial vaccination could be crucial in highlighting the role of vaccines as an effective tool in fight against the COVID-19 during these times of vaccine misinformation to motivate more people to get vaccinated. We found 11% patients with RT-PCR negative but with classic COVID-19 symptoms which we think could be due to false negative tests, stage of the disease where nasopharyngeal swab is inconclusive or the newer variant of the virus^14 15^.

Our study has several limitations being a retrospective single center study. We did not have patient details other than their basic demographics, COVID-19 RT-PCR status, vaccination status and CT severity score. Patient details such as their oxygen saturations, hospital admission status, co-morbidities and mortality were not available.

In conclusion, we present here the real-world findings from an independent radiology practice (a unique and common practice model), in India amid the second COVID-19 wave showing significantly lower CT severity score in fully or partially vaccinated patients compared to unvaccinated patients. We report a higher CT severity score in patients with positive RT-PCR. Complete vaccination in patients could be critical in preventing severe lung disease.

## Data Availability

The de-identified data maybe available upon request. Subject to approval of local ethical committee.

## Statements

### Contributorship Statement

Drs. Revat Lakhia and Jaimin Trivedi contributed equally in design, conceptualization, administration and writing of the manuscript. Dr. Lakhia was lead data capturer, curator and manager. Dr. Trivedi was lead data analyst.

### Ethical Committee Statement

This work was approved by an independent ethical committer at the ACEAS in Ahmedabad, India, registered with the OHRP US DHHS Reg No. IRB00011046.

